# Excess mortality during the 1918–20 influenza pandemic in Czechia

**DOI:** 10.1101/2021.01.10.21249537

**Authors:** Hampton Gray Gaddy

## Abstract

This research letter provides a replicable estimate of the mortality that the 1918–20 influenza pandemic caused in Czechia. A monthly all-cause excess mortality model identified clear periods of pandemic mortality in September 1918 through May 1919 and January 1920 through May 1920. The total excess mortality in those months implies a pandemic death toll of 71,967 and a national death rate of 0.75%.

## Introduction

Given its near-global scope and short duration, the influenza pandemic that began in 1918 was one of the highest mortality events in modern human history. Since the resumption of interest in the pandemic in the 1990s, global, national, and local literatures about the pandemic have steadily grown (1–4). This research letter estimates the mortality that the pandemic caused in the Central European country of Czechia.

In general, the pandemic caused mortality over several waves. In parts of the world with reliable data, most deaths occurred in two successive waves during the last quarter of 1918 and first quarter of 1919. For 14 national European populations, roughly 0.34% (Denmark) to 2.23% (Portugal) of each population died in those two waves (4). Defining the temporal boundaries of the pandemic is difficult (5–7), but a clinically similar wave in early 1920 (4) and a clinically less severe, ‘herald’ wave earlier on in 1918 are also well documented (8).

Cause of death attribution in the face of a poorly understood threat is often difficult. As such, those outlying waves were substantiated and the mortality of the main waves themselves is often estimated using excess mortality models (4, 5, 7, 9). These models assume an all-cause or cause-specific baseline mortality rate for a population and apply it as a counterfactual to the observed mortality data from the pandemic period. The difference between the observed and counterfactual mortality is taken as the pandemic death toll, subject to caveats about the validity of the counterfactual (10) and how proximate and ultimate causes of death are conceptualized (11–14).

## Methods

The Czech Statistical Office reports monthly all-cause death counts for the Czech lands — the territory that comprises the Czech parts of Austria-Hungary, the Czech part of Czechoslovakia, and the modern state of Czechia. The monthly counts from January 1914 to December 1923 (15, 16) were fitted to a cyclical Serfling regression model that estimated temporal and seasonal fluctuation in baseline mortality over the study period (9, 17). The months of October and November 1918 and February and March 1920 were removed from the training set, as these were the months with the most abnormal mortality by far. For all periods historically associated with the pandemic, months of all-cause deaths that were significantly in excess of the baseline regression fit were interpreted as periods of pandemic mortality. My results are fully replicable with the provided death count data and R code (18).

## Results

Figure 1 displays the observed monthly death totals in Czechia between 1914 and 1923. It also shows the 99.9% confidence interval of the fitted cyclical regression model as an upper limit on baseline mortality. There is clear excess mortality in the periods of September 1918 through May 1919 and January 1920 through May 1920. The total difference between the observed and mean baseline mortality in those months is 71,967 deaths. Out of an estimated 1918 population of 9,624,230 in the Czech lands (15), this implies a pandemic death rate of 0.75%.

**Figure 1.**
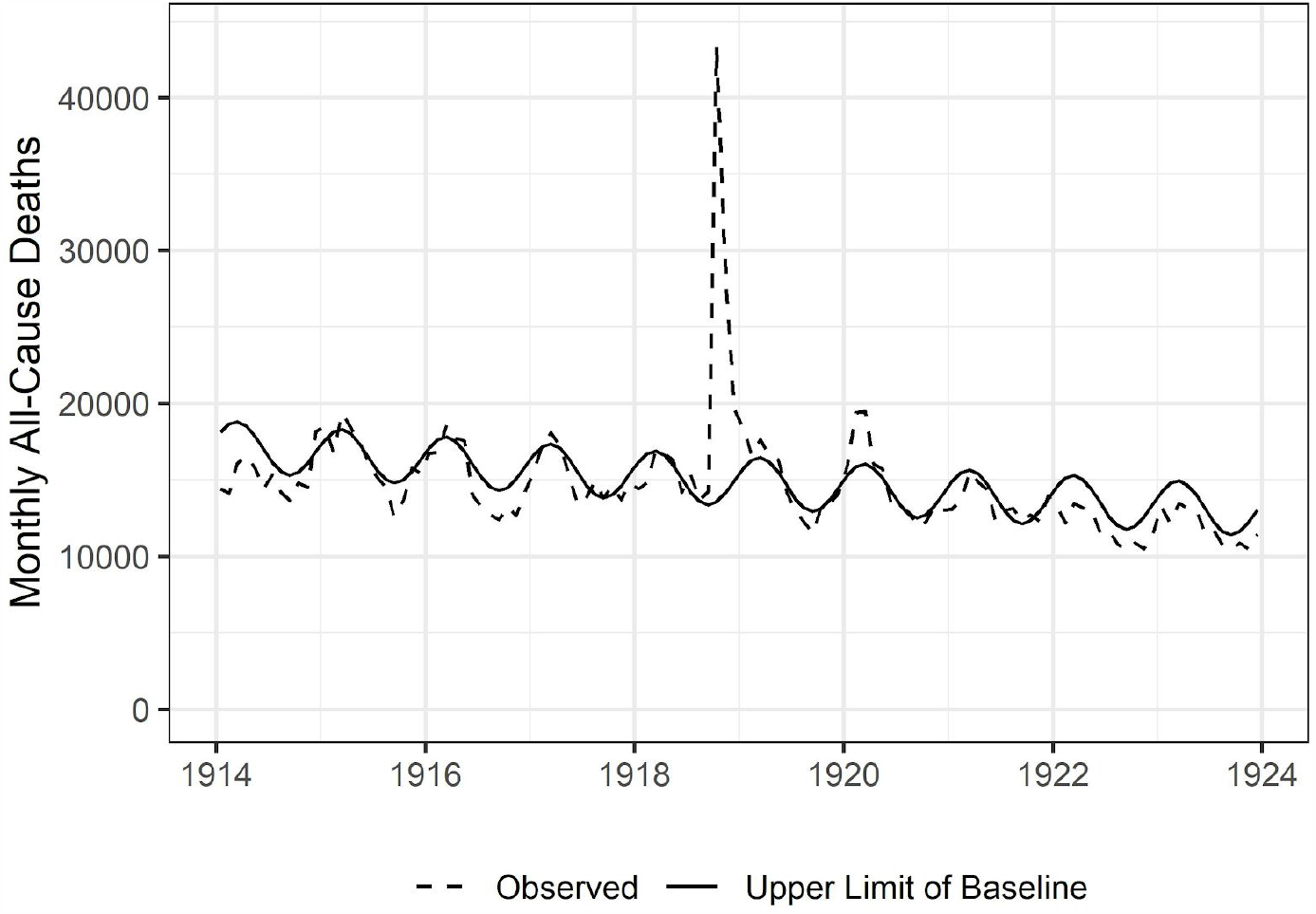

The model suggests that there were 51,763 deaths (72% of the total) between September and December 1918, 8,526 deaths (12% of the total) between January and May 1919, and 11,678 deaths (16% of the total) between January and May 1920. There is less clear evidence for pandemic mortality before September 1918. Observed mortality in May and July 1918 did clearly exceed the confidence interval on the baseline mortality regression. Those months saw 950 and 1,719 excess deaths relative to the mean baseline estimate. The existence of herald mortality in May 1918 in Central Europe is not generally supported by the literature, but the much more reasonable inclusion of excess mortality from July 1918 increases the total pandemic death toll to 73,686. This yields an estimated total pandemic death rate of 0.77%.

## Discussion

In terms of mortality, Czechia had a seemingly similar experience of the 1918–20 pandemic to other states in Western and Central Europe. For the waves at the end and beginning of 1918 and 1919, respectively, I calculate an excess death toll of 60,289 and an excess death rate of 0.63%. This is similar to the monthly all-cause excess death rates of the Netherlands (0.62%), Germany (0.66%), France (0.73%), and Switzerland (0.77%) in those waves (4).

To my knowledge, this report is the first formal mortality study of the 1918–20 pandemic in Czechia, but it is preceded by a significant dissertation published in 2017 (19). As part of a historical overview of the outbreak in Czechia, it performed five rough mortality calculations that placed the national death toll at between 44,000 and 77,000. My estimate suggests that the true death toll fell on the higher end of that range.

This analysis has several limitations. Monthly death counts are assumed to be correct, and in particular, it is assumed that death registration did not vary non-linearly over the course of the study period. Death registration could have been relatively incomplete in the autumn of 1918, in the face of the contemporary pandemic and political regime changes. If so, this would suggest a higher death toll than the one I report. There is also a discontinuity in the source of the monthly death count data between 1918 and 1919 (15, 16), but this seems to have no effect on the baseline trend (Figure 1). The estimation of a pandemic death rate assumes the accuracy of the national population count used. The population count used does not include military forces (19), and it is assumed that monthly death counts did not include them either. An estimated 5,000 to 10,000 additional pandemic deaths are estimated for the military, out of an unknown at-risk population (19). The standard assumption about the validity of excess mortality counterfactuals also applies.

However, this estimate was conducted in a similar manner to most other European mortality estimates for the 1918–20 pandemic. It strengthens and broadens the growing consensus on the mortality that Europe experienced in that pandemic.

## Data Availability

All data and methods used are available via links referenced in the manuscript. The source data (.csv) and code (.R) used are also provided.

