## Supplementary material for "Excess mortality during the 1918–20 influenza pandemic in Czechia": Data and code: czechia_spanishflu.R

```
#This code is licensed under GPL-3 (https://choosealicense.com/licenses/gpl-3.0/)

#load packages
#all packages used under GPL licenses
if (!require("pacman")) install.packages("pacman")
library(pacman)
p_load("devtools", "tidyverse")
devtools::install_github("kmcconeghy/flumodelr")
library(flumodelr)

#load data
#data used and distributed under the terms of the Czech Statistical Office (https://web.archive.org/web/20201119091923/https://www.czso.cz/csu/czso/podminky_pro_vyuzivani_a_dalsi_zverejnovani_statistickych_udaju_csu)
data = read.csv("czechia_spanishflu.csv", fileEncoding="UTF-8-BOM")
data$time = as.Date(data$time, format="%Y-%m-%d")

#include all months in model except 10/18, 11/18, 2/20, 3/20
data$epi = FALSE
data$epi[58] = TRUE
data$epi[59] = TRUE
data$epi[74] = TRUE
data$epi[75] = TRUE

#model
fit = fluserf(data=data, outc=deaths, epi=epi, time=time, period=12, alpha=0.001)
colnames(fit) = c("year", "month", "time", "Observed", "epi", "Baseline", "Upper Limit of Baseline")

#figure 1
lty = c("Observed"="dashed", "Upper Limit of Baseline"="solid")
ggplot(data=fit) +
  geom_line(aes(time, Observed, lty="Observed")) +
  geom_line(aes(time, `Upper Limit of Baseline`, lty="Upper Limit of Baseline")) +
  scale_linetype_manual(values=c("dashed", "solid"), limits=c("Observed", "Upper Limit of Baseline")) +
  lims(y=c(0, 44000)) +
  labs(x=NULL, y="Monthly All-Cause Deaths", linetype=NULL) +
  theme_bw() +
  theme(legend.position="bottom", plot.title = element_text(hjust = 0.5)) +
  ggtitle("Figure 1")

#save figure 1
ggsave("Fig1.jpeg", dpi=500, scale=1.25)

#death toll of main 1918-1919 waves + 1920 wave
round(sum(fit$Observed[57:65]-fit$Baseline[57:65]) + sum(fit$Observed[73:77]-fit$Baseline[73:77]))

#death toll including potential herald deaths in 7/18
round(sum(fit$Observed[57:65]-fit$Baseline[57:65]) + sum(fit$Observed[73:77]-fit$Baseline[73:77]) + fit$Observed[55]-fit$Baseline[55])
```
